# Combining Clinical LAFOV PET/CT with a Digital Twin Providing Motion-Free Ground Truth Reveals Quantitative Trade-offs in Respiratory Motion Correction

**DOI:** 10.64898/2026.08.11.26360175

**Authors:** W. Lan, S. Weigel, E. Calderón, C. la Fougère, F. P. Schmidt

**Author notes:** Equal contribution (W. Lan, S. Weigel).

## Abstract

**Purpose:** Respiratory motion remains a major source of quantitative bias in PET and becomes increasingly relevant for high-sensitivity long axial field-of-view (LAFOV) PET/CT. Although numerous respiratory motion correction (MoCo) methods have been proposed, their quantitative accuracy cannot be established clinically because a patient-specific motion-free reference is fundamentally unavailable in vivo. This study combined clinical PET imaging with a digital twin, a realistic representation of both the PET/CT system and the patient, to objectively validate respiratory MoCo against a corresponding motion-free reference.

**Methods:** Twenty patients (10 [¹⁸F]FDG with predominantly pulmonary lesions and 10 [¹⁸F]SiFA*lin*-TATE with predominantly hepatic lesions; total 135 lesions) were analyzed. The digital twin combined a validated LAFOV PET/CT simulation model with an anatomically realistic phantom containing 14 lung and liver lesions, two patient-derived respiratory patterns, and respiratory motion amplitudes of 2 and 3 cm, generating patient-like datasets with corresponding motion-free references. Data-driven and image-based MoCo were evaluated using lesion morphology, SUV_mean_, SUV_max_, and metabolic tumor volume (MTV).

**Results:** In patients, data-driven MoCo produced larger SUV_mean_ increases than image-based MoCo for liver (48.1±18.9% vs. 17.0±12.0%; p<0.01), lower-lung (32.5±21.2% vs. 16.3±15.6%, p=0.06), and upper-lung lesions (28.4±32.0% vs. 10.4±17.2%; p<0.01), with similar findings for SUV_max_ and larger MTV reductions. Simulation revealed marked motion-induced SUV_mean_ underestimation before correction, particularly in liver (−31.2±6.8%) and lower lung (−15.5±13.9%). Relative to the motion-free reference, data-driven MoCo most accurately recovered hepatic uptake (4.3±11.7% vs. −10.0±9.2%; p=0.01) but overestimated pulmonary uptake (lower lung: 19.8±16.3% vs. −1.6±10.2%; p=0.02). SUV_max_ showed the same regional behavior, whereas image-based MoCo yielded MTV estimates closer to the reference. Quantitative recovery was largely independent of respiratory pattern, while larger motion amplitudes mainly affected image-based MoCo.

**Conclusion:** Combining clinical PET with a realistic digital twin and corresponding motion-free ground truth enabled objective validation of respiratory MoCo beyond conventional clinical evaluation. Larger correction-induced quantitative changes should not be equated with greater quantitative accuracy. Instead, MoCo performance was region- and metric-dependent, highlighting the value of ground-truth-based validation for developing and benchmarking respiratory motion correction and quantitative PET on LAFOV PET/CT systems.

## Introduction

Respiratory motion remains a major source of quantitative error in PET/CT. During free-breathing acquisitions, respiratory displacement causes lesion blurring, PET/CT misregistration, SUV underestimation, and metabolic tumor volume (MTV) overestimation, particularly for lesions adjacent to the diaphragm. These motion-induced quantitative biases may adversely affect lesion characterization, therapy-response assessment, radiotherapy planning, and quantitative analyses (*1*,*2*).

The advent of long-axial-field-of-view (LAFOV) PET/CT has further increased the importance of accurate motion management. Owing to their markedly improved sensitivity, LAFOV systems enable high-temporal-resolution dynamic imaging, and whole-body parametric imaging with unprecedented quantitative precision (*3*). As statistical uncertainty decreases, physiological motion is emerging as one of the major remaining barriers to accurate PET quantification, a challenge expected to become even more relevant as LAFOV PET expands towards more advanced quantitative imaging applications (*4*,*5*).

Several respiratory motion correction strategies have been developed for PET, including sensor-based (*6*) and device-less approaches (*7*) that estimate respiratory motion either directly from PET acquisition data (*8*) or from reconstructed images (*9*,*10*). As associated methods operate at different stages of the PET processing chain, differences in quantitative recovery, lesion delineation, and image noise are expected (*11*).

Despite these methodological differences, respiratory motion correction is typically evaluated by comparison with conventional static reconstructions (*12*,*13*). Such comparisons quantify correction-induced changes but cannot determine quantitative accuracy because a patient-specific motion-free reference is inherently unavailable in vivo. Overcoming this fundamental limitation requires a framework capable of generating patient-like datasets with and without respiratory motion under otherwise identical imaging conditions. We recently developed and validated a scanner-specific digital twin of the Biograph Vision Quadra LAFOV PET/CT system that combines a realistic patient-like phantom capable of modelling respiratory motion (*14*) with Monte Carlo PET simulation (*15*) reproducing the complete clinical acquisition and image reconstruction workflow (*16*).

To our knowledge, this is the first study to combine clinical evaluation with a validated patient-like digital twin to objectively benchmark respiratory motion correction against a corresponding motion-free reference.

We hypothesized that benchmarking against such a reference would identify the correction strategy that most accurately recovers the underlying tracer distribution while quantifying the residual motion-induced errors that remain after correction. Furthermore, we hypothesized that greater apparent recovery of quantitative PET metrics does not necessarily reflect higher quantitative accuracy, as stronger motion correction may also introduce overcompensation.

Accordingly, this study combined clinical and simulation-based evaluations of respiratory motion correction in LAFOV PET/CT. Clinical data quantified the effects of data-driven and image-based motion corrections on lesion uptake, metabolic tumor volume, and image quality, whereas the digital twin enabled objective assessment of quantitative accuracy against a corresponding motion-free reference.

## Materials and Methods

### Clinical Datasets

Two complementary clinical cohorts (n = 10 each) were included: an [¹⁸F]FDG cohort with predominantly pulmonary lesions and an [¹⁸F]SiFA*lin*-TATE cohort with predominantly hepatic lesions, representing distinct tracer distributions and respiratory motion characteristics. The [¹⁸F]SiFA*lin*-TATE cohort comprised patients with somatostatin receptor-expressing neuroendocrine tumors (age: 61±8 y; weight: 89±18 kg; injected activity: 191±35 MBq), whereas the [¹⁸F]FDG cohort consisted of patients with lung cancer (age: 70±8 y; weight: 78±11 kg; injected activity: 156±21 MBq). The study was approved by the institutional review board (773/2021BO2) and written informed consent was obtained from all patients.

### PET Acquisition and Image Reconstruction

Clinical PET/CT examinations were performed on a Biograph Vision Quadra LAFOV PET/CT system (Siemens Healthineers, Knoxville, TN, USA). Patients underwent 5-min whole-body acquisitions according to the respective clinical imaging protocols. All PET datasets were reconstructed using the vendor’s investigational prototype e7 tools (OP-OSEM, 4 iterations, 5 subsets, PSF, TOF, matrix size 440×440, voxel size 1.65×1.65×1.65 mm^3^). The identical reconstruction pipeline was implemented within the digital twin framework.

### Patient-like Digital Twin Framework

A previously developed and validated digital twin of the Biograph Vision Quadra LAFOV PET/CT system served as the simulation framework for this study (*16*). A 50th-percentile adult female XCAT phantom (*14*) was used to simulate a 5-min whole-body [¹⁸F]FDG PET acquisition. Fourteen spherical lesions (diameters: 10 and 15 mm; activity concentrations: 7, 14, and 28 kBq/mL) were placed in the liver and lungs (**FIGURE. 1A**). Lesion size, activity concentration, and anatomical distribution were selected to reflect the characteristics observed in the clinical cohorts.

**FIGURE. 1.**
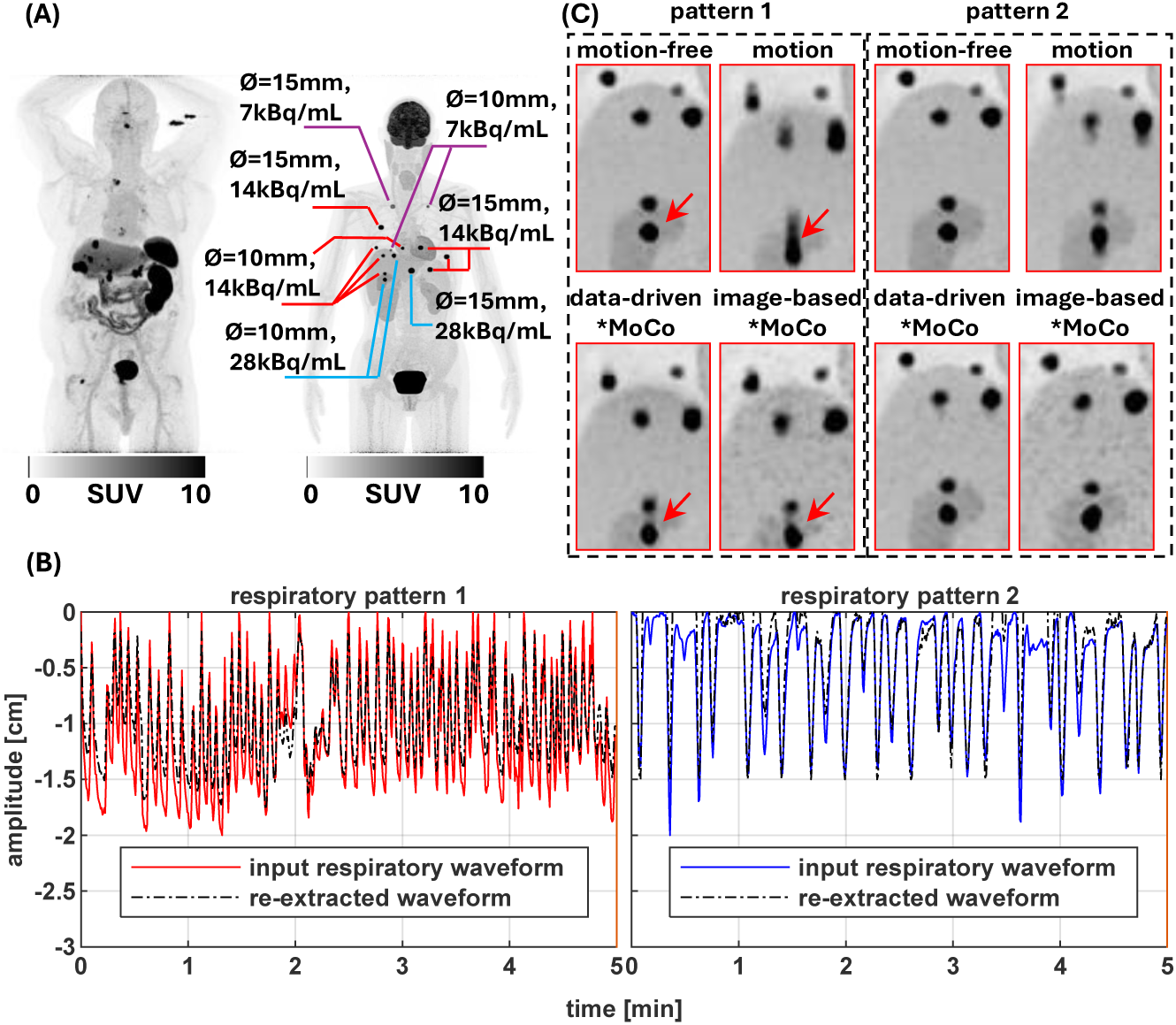
Patient-like digital twin framework used for objective benchmarking of respiratory motion correction (MoCo). (A) Comparison of a representative clinical PET maximum-intensity projection (left) and the corresponding patient-like XCAT-based digital phantom (right), illustrating the close similarity between clinical imaging and the simulated patient model. Simulated liver and lung lesions with clinically representative diameters and activity concentrations are indicated. (B) Two patient-derived respiratory patterns (pattern 1 and 2). Solid lines represent the respiratory waveforms prescribed as input to the simulation, whereas dashed lines show the corresponding waveforms re-extracted from the simulated PET data using the same data-driven respiratory gating algorithm, demonstrating accurate implementation of the respiratory motion model. (C) Representative simulated PET images for both respiratory patterns showing the motion-free reference, respiratory motion, and the corresponding data-driven and image-based MoCo results.

Respiratory motion was modeled using two patient-derived respiratory waveforms (pattern-1 and pattern-2) extracted from clinical PET data with the commercially available OncoFreeze AI (Siemens Healthineers, Knoxville, TN, USA) (*8*,*17*) and incorporated into the XCAT phantom with amplitudes of 2 and 3 cm. Accurate implementation was confirmed by re-extracting the respiratory waveforms from the simulated PET data (**FIGURE. 1B**). Corresponding motion-free datasets served as the reference for residual error quantification (**FIGURE. 1C**).

### Respiratory Motion Correction

Two fundamentally different respiratory motion correction (MoCo) strategies were evaluated: a data-driven approach integrated into image reconstruction and an image-based approach applied after image reconstruction (**FIGURE. 1C**). Data-driven MoCo was performed using OncoFreeze AI (*8*,*17*) (**FIGURE. 2A**), which estimates respiratory motion directly from PET listmode data and incorporates voxel-wise motion vector fields into iterative reconstruction. Image-based MoCo was implemented using an adapted version of the Fast Algorithm for Motion Correction (FALCON, Medical University of Vienna, Vienna, Austria) (*9*). Static PET listmode data were rebinned into consecutive 2-s frames, non-rigidly registered to a reference frame, and averaged to generate the final motion-corrected image (**FIGURE. 2B**).

**FIGURE. 2.**
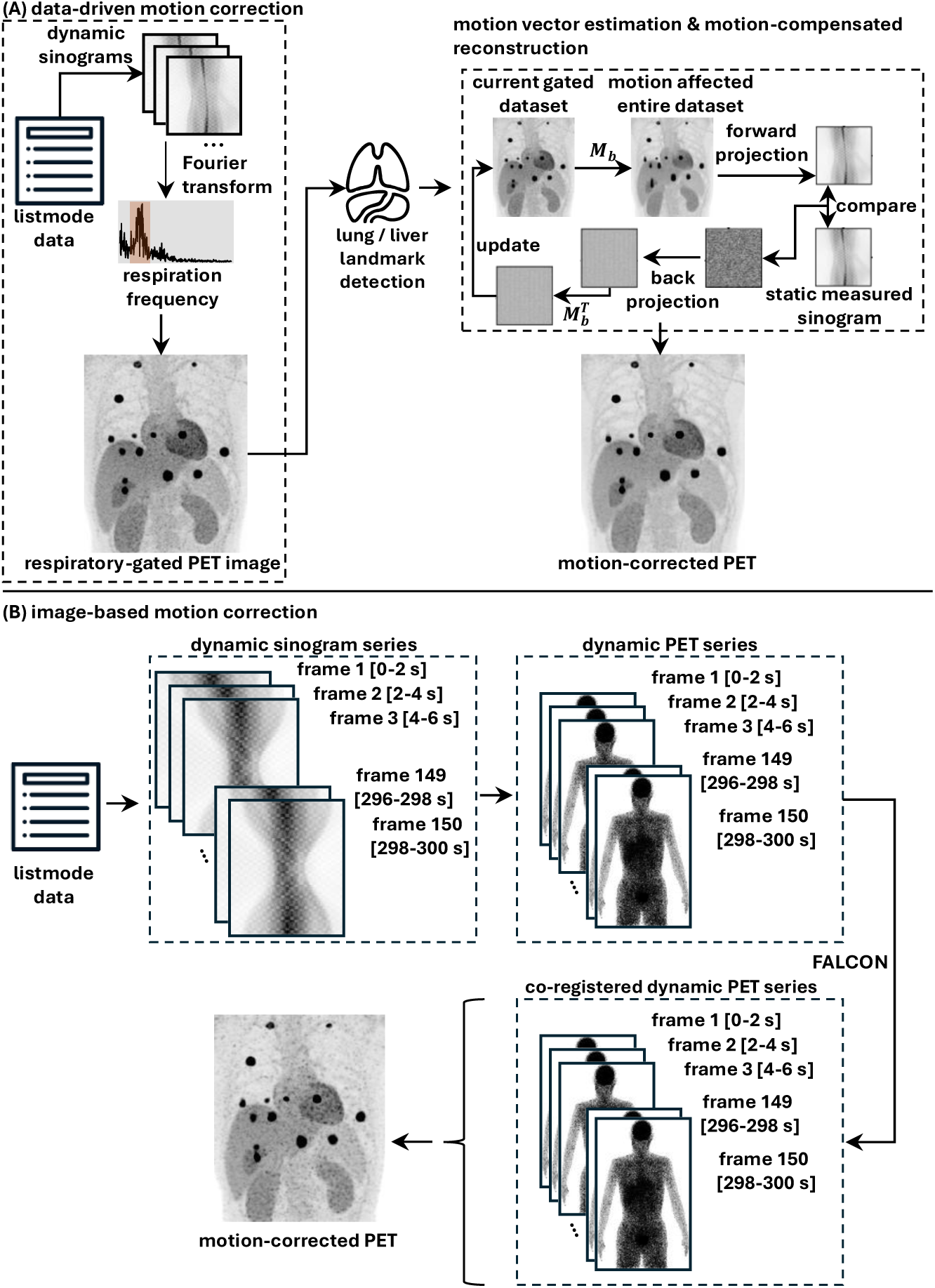
Processing workflows of the two respiratory motion correction (MoCo) strategies. (A) Data-driven MoCo using OncoFreeze AI. A respiratory-gated PET image was first reconstructed, followed by automatic lung and liver landmark detection and estimation of voxel-wise motion vector fields, which were incorporated into the iterative reconstruction to generate the final motion-corrected PET image. (B) Adapted image-based MoCo using the Fast Algorithm for Motion Correction (FALCON). Static PET listmode data were converted into a dynamic image series by rebinning into consecutive 2-s frames. The reconstructed image series was non-rigidly registered to a reference frame using multiscale greedy diffeomorphic image-registration, followed by averaging of the registered frames to generate the final motion-corrected static PET image.

### Quantitative Lesion Segmentation and Analysis

The same quantitative analysis pipeline was applied to clinical and simulated datasets. Lesions were segmented using a 50% SUV_max_ threshold in Hermes Affinity (Version 5.0.1; Hermes Medical Solutions) and 3D Slicer (Version 5.8.2) (*18*). Segmentation was performed independently for uncorrected, motion-corrected, and motion-free reference datasets. SUV_mean_, SUV_max_, and metabolic tumor volume (MTV) were extracted, and lesions were categorized according to their anatomical location as upper lung, lower lung, or liver.

For the clinical datasets, respiratory motion correction was evaluated as the relative change compared with the corresponding uncorrected images. To avoid overweighting patients with multiple lesions, lesion measurements were averaged at the patient level within each anatomical region before statistical analysis. For the simulated datasets, residual quantitative error was calculated as the signed percentage difference between motion-corrected and motion-free reference values. Negative values indicate underestimation and positive values overestimation relative to the motion-free reference.

Image noise was quantified using the coefficient of variation (CV), calculated as the ratio of the standard deviation to the mean activity concentration measured within a 30-mm spherical volume of interest placed in homogeneous liver tissue.

### Statistical Analysis

Normality was assessed using the Shapiro–Wilk test. Paired t-tests or Wilcoxon signed-rank tests were used for paired method comparisons. The effects of respiratory pattern and motion amplitude were assessed using paired lesion-level comparisons of the signed percentage error relative to the motion-free reference. Respiratory-pattern effects were assessed after averaging across both motion amplitudes, whereas motion-amplitude effects were assessed after averaging across both respiratory patterns. All statistical tests were two-sided, and p values < 0.05 were considered statistically significant. Statistical analyses were performed using MATLAB R2026a (MathWorks, Natick, MA, USA).

## Results

### Clinical Impact of Respiratory Motion Correction

Representative examples of respiratory motion correction are shown in **FIGURE. 3**. Respiratory motion resulted in visible lesion blurring, particularly for hepatic and peripheral pulmonary lesions adjacent to the diaphragm. Both MoCo strategies reduced motion-induced blurring, with the data-driven approach producing sharper lesion delineation than the image-based approach.

**FIGURE. 3.**
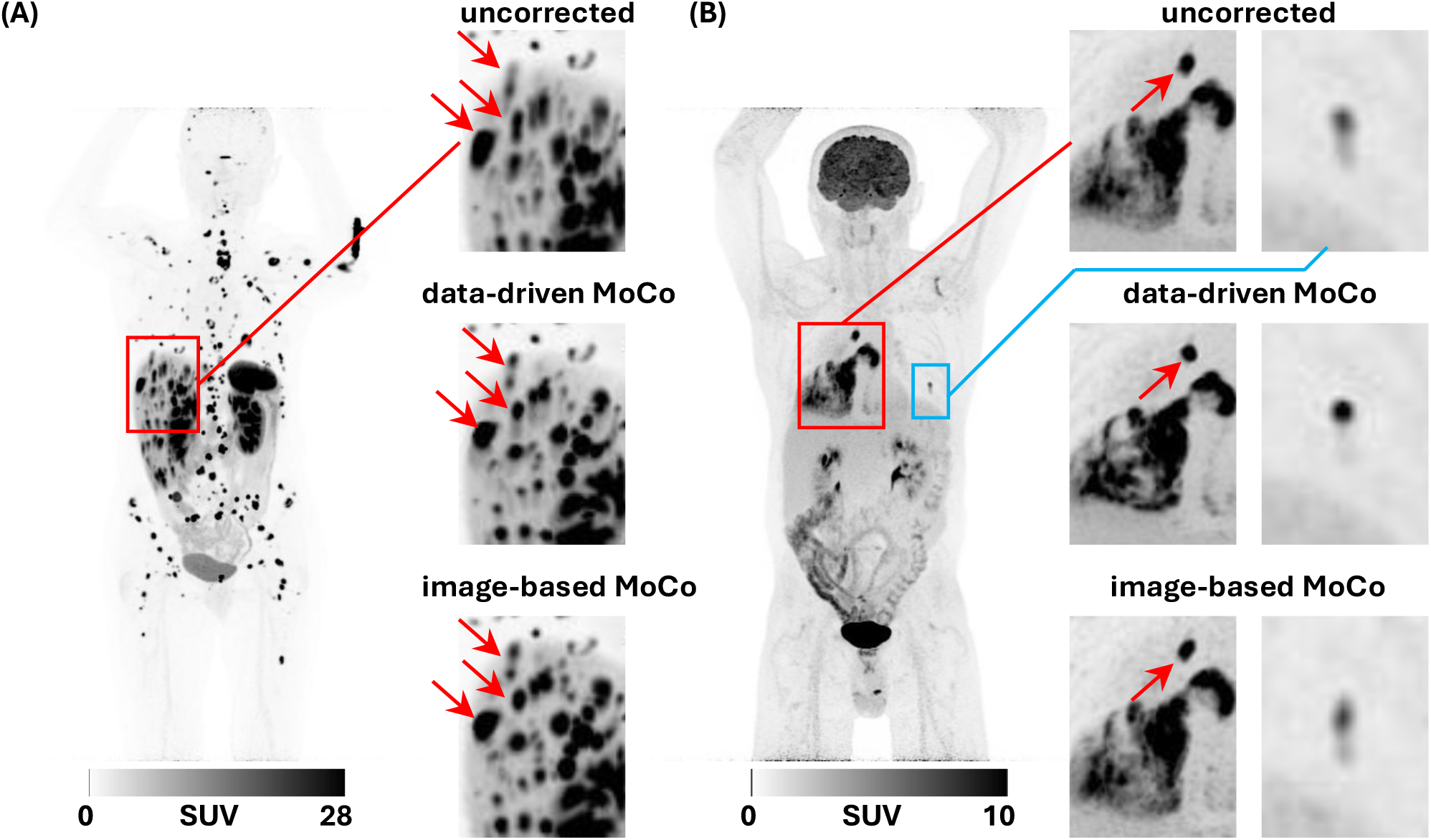
Representative clinical examples of respiratory motion correction. Maximum-intensity projections (MIPs) and enlarged lesion views from (A) [¹⁸F]SiFA*lin*-TATE and (B) [¹⁸F]FDG PET/CT. Both data-driven and image-based motion correction (MoCo) reduce motion-induced lesion blurring. Arrows indicate representative morphological changes.

Image noise remained largely unchanged (**FIGURE. 3**), with CV increasing from 9.5 ± 1.5% in the uncorrected images to 10.3 ± 1.6% following data-driven and 11.4 ± 2.0% following image-based MoCo.

Quantitative analysis included 135 lesions from 20 patients (71 liver, 36 lower-lung, and 28 upper-lung lesions). The largest correction-induced changes were consistently observed for liver lesions, where data-driven MoCo resulted in substantially larger increases in SUV_mean_ than image-based MoCo (48.1 ± 18.9% vs. 17.0 ± 12.0%, p < 0.01, **FIGURE. 4-1**).

**FIGURE. 4.**
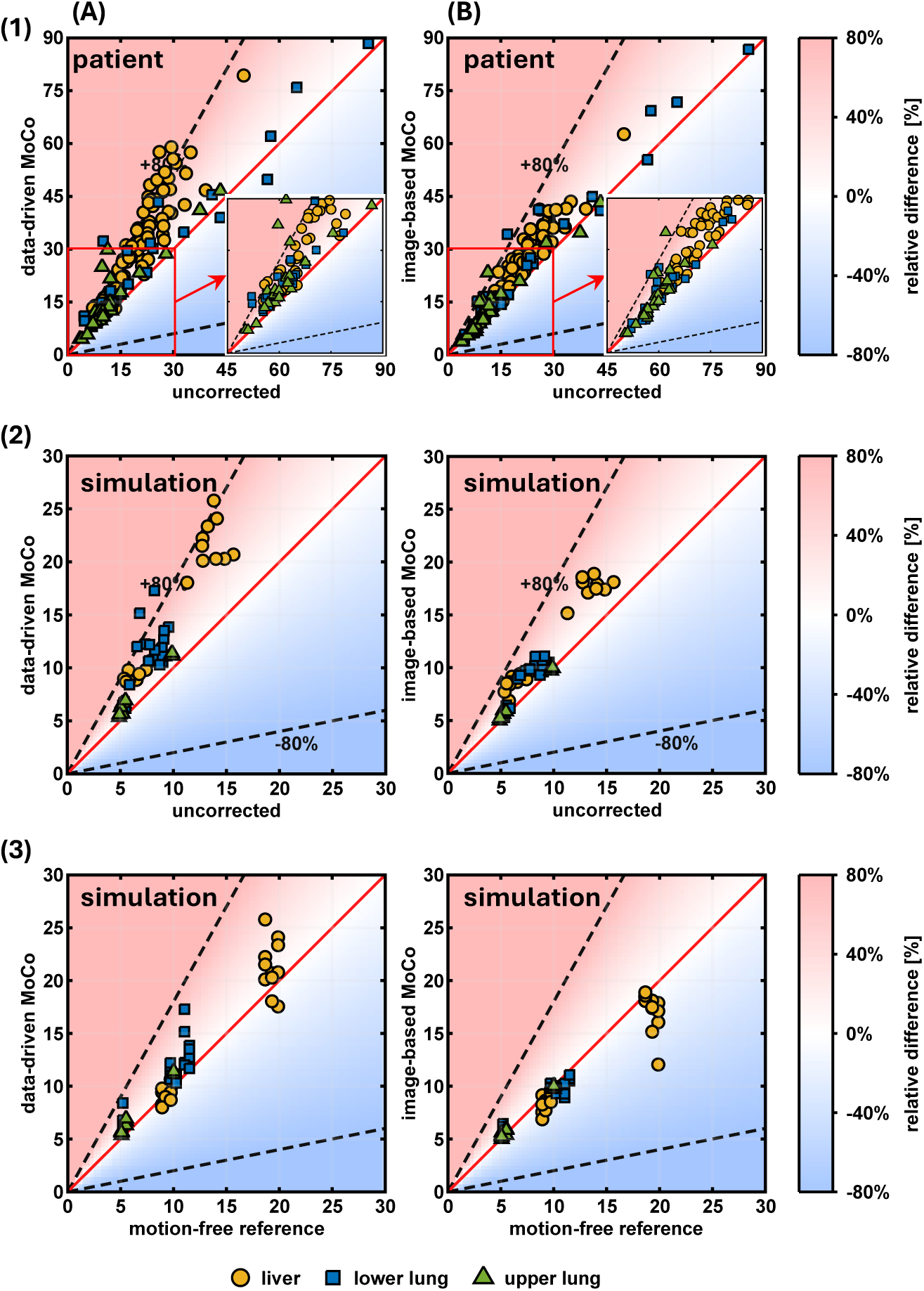
Quantitative comparison of SUV_mean_. Scatter plots compare lesion SUV_mean_ values after respiratory motion correction with the corresponding uncorrected or motion-free reference values for clinical and simulated datasets. (1) Clinical datasets showing correction-induced changes after data-driven (A) and image-based (B) MoCo relative to uncorrected images. Insets highlight lesions with SUV_mean_ between 10 and 30. (2) Patient-like digital twin demonstrating close agreement with the regional correction patterns observed in the clinical datasets under identical respiratory motion conditions. (3) Comparison of the simulated motion-corrected datasets with the corresponding motion-free reference, after data-driven (A) and image-based (B) MoCo. Circles, squares, and triangles denote liver, lower-lung, and upper-lung lesions.

A similar trend was observed for lower-lung lesions (32.5 ± 21.2% vs. 16.3 ± 15.6%, p = 0.06), whereas upper-lung lesions showed smaller changes (28.4 ± 32.0% vs. 10.4 ± 17.2%, p < 0.01).

Comparable findings were observed for SUV_max_ (**Supplemental Figure 1**). MTV consistently decreased to a greater extent following data-driven than image-based MoCo (**FIGURE. 5-1**), consistent with a more compact lesion representation. Complete quantitative results are provided in **Supplemental Table 1**.

**FIGURE. 5.**
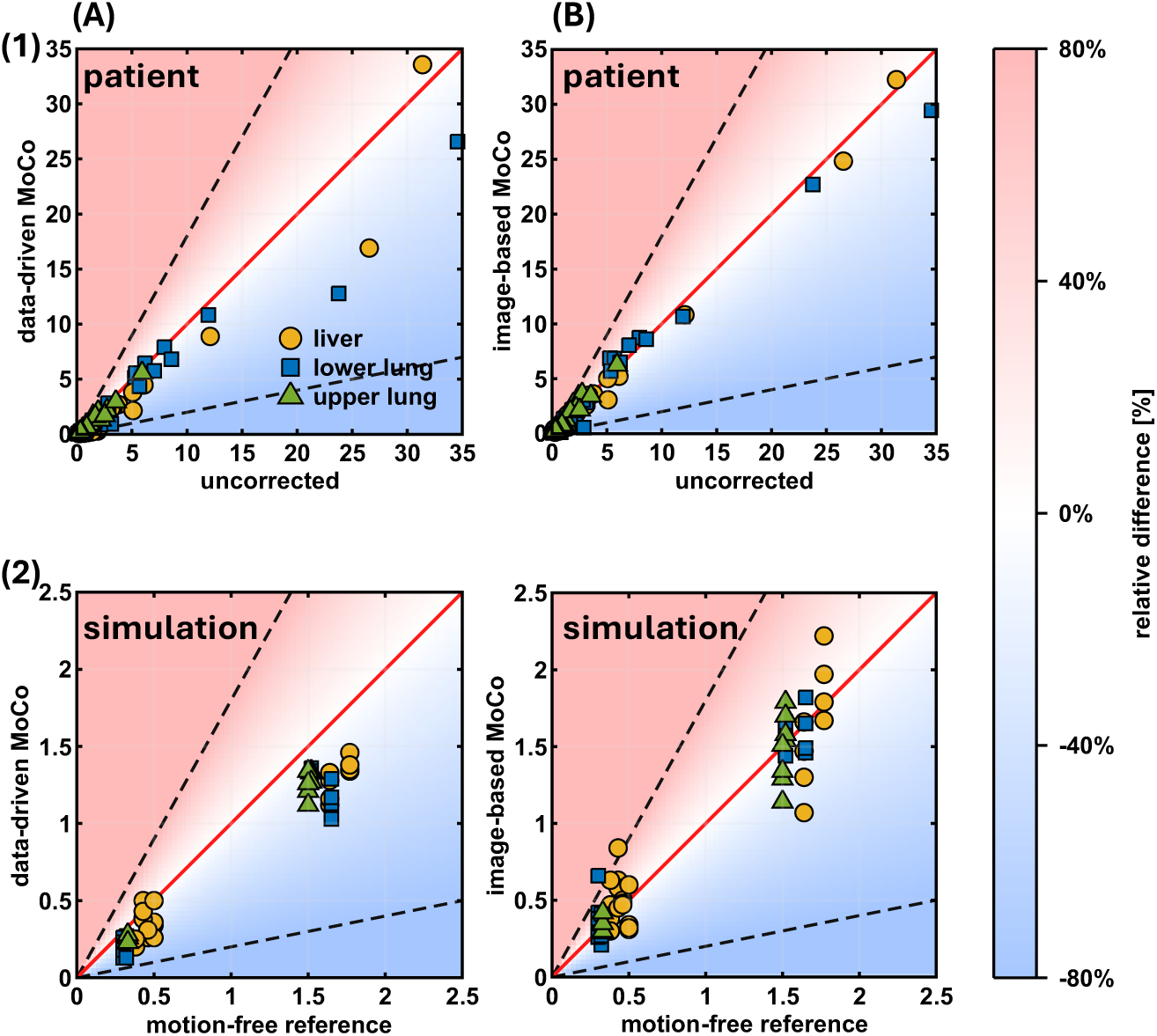
Quantitative comparison of MTV. Scatter plots compare lesion MTV values after data-driven (A) and image-based (B) MoCo with the corresponding uncorrected values in the clinical datasets (1) and the corresponding motion-free reference in the simulated datasets (2). Circles, squares, and triangles denote liver, lower-lung, and upper-lung lesions.

### Simulation-Based Validation of Clinical Findings

The patient-like digital twin closely reproduced the qualitative and quantitative findings observed in the clinical datasets (**FIGURE. 1C**). Similar to the clinical observations, the data-driven MoCo produced more compact lesion appearance and sharper lesion boundaries than the image-based MoCo.

Image noise showed the same behavior, with the CV remaining nearly unchanged for the motion-free reference (8.8%), uncorrected images (8.7 ± 0.1%), and data-driven MoCo (9.5 ± 0.4%), while image-based MoCo resulted in the highest image noise (11.9 ± 0.5%).

Quantitative analysis likewise mirrored the clinical findings. The largest correction-induced changes were observed for liver lesions, followed by lower-lung lesions, whereas upper-lung lesions showed only minor changes. Data-driven MoCo consistently produced larger changes than image-based MoCo for SUV_mean_ (**FIGURE. 4**), MTV (**FIGURE. 5**) and SUV_max_ (**Supplemental Figure 1**). For liver lesions, SUV_mean_ increased by 55.3 ± 16.1% following data-driven MoCo compared with 34.5 ± 11.7% following image-based MoCo. Similar trends were observed for lower-lung lesions (45.1 ± 29.3% vs. 18.0 ± 11.3%), whereas upper-lung lesions exhibited only minor changes. Complete quantitative results are summarized in **Supplemental Table 1**.

### Residual Quantitative Error Relative to the Motion-Free Reference

The availability of a corresponding motion-free reference enabled objective assessment of whether correction-induced changes reflected improved quantitative recovery.

Relative to the motion-free reference, respiratory motion introduced substantial quantitative errors that were strongly location dependent. The largest SUV underestimation occurred in liver lesions, where SUV_mean_ was reduced by 31.2 ± 6.8%, followed by lower-lung lesions (15.5 ± 13.9%), whereas upper-lung lesions were only minimally affected (−2.5 ± 2.7%) (**FIGURE. 4-3**).

Both motion correction strategies markedly reduced these motion-induced errors. For liver lesions, data-driven MoCo restored SUV_mean_ from −31.2 ± 6.8% to 4.3 ± 11.7%, whereas image-based MoCo reduced the error to −10.0 ± 9.2% (p = 0.01), indicating that the stronger correction achieved by the data-driven approach resulted in quantitative values closer to the motion-free reference. In contrast, for lower- and upper-lung lesions, data-driven MoCo tended to overcorrect, yielding SUV_mean_ errors of 19.8 ± 16.3% and 14.5 ± 5.3%, respectively, whereas image-based MoCo remained closer to the motion-free reference (−1.6 ± 10.2% and 3.0 ± 4.4%). Similar regional behavior was observed for SUV_max_ (**Supplemental Figure 1**), demonstrating that the relative performance of both correction strategies depended on lesion location.

MTV exhibited the opposite trend. Respiratory motion overestimated lesion volume, particularly for liver lesions (57.4 ± 82.5%) and lower-lung lesions (14.0 ± 26.6%). Image-based MoCo produced MTV values closer to the motion-free reference but with considerably larger variability, whereas data-driven MoCo yielded more consistent recovery (standard deviations of 15.6% and 13.6% for liver and lower-lung lesions, respectively), despite a tendency towards overcorrection (mean errors of −25.2% and −26.6%). Complete quantitative results are summarized in **TABLE. 1**.

**TABLE. 1.** Relative quantitative error of simulated lesions with respect to the motion-free reference. Mean signed percentage differences (±SD) for uncorrected, data-driven, and image-based MoCo are shown separately for liver, lower-lung, and upper-lung lesions. Positive values indicate overestimation and negative values underestimation relative to the motion-free reference.

| parameter | structure | uncorrected | data-driven MoCo | image-based MoCo | p-value* |
| --- | --- | --- | --- | --- | --- |
| SUV<sub>mean</sub><br>(phantom) | liver | -31.2 ± 6.8 | 4.3 ± 11.7 | -10.0 ± 9.2 | 0.01 |
| | lower lung | -15.5 ± 13.9 | 19.8 ± 16.3 | -1.6 ± 10.2 | 0.02 |
| | upper lung | -2.5 ± 2.7 | 14.5 ± 5.3 | 3.0 ± 4.4 | 0.06 |
| SUV<sub>max</sub><br>(phantom) | liver | -27.2 ± 7.7 | 8.6 ± 13.1 | -4.8 ± 10.6 | 0.02 |
| | lower lung | -11.8 ± 14.3 | 24.0 ± 17.3 | 2.7 ± 11.0 | 0.01 |
| | upper lung | -2.5 ± 5.3 | 18.2 ± 5.0 | 8.0 ± 9.7 | 0.12 |
| MTV<br>(phantom) | liver | 57.4 ± 82.5 | -25.2 ± 15.6 | 6.6 ± 31.4 | 0.01 |
| | lower lung | 14.0 ± 26.6 | -26.6 ± 13.6 | 8.3 ± 31.1 | 0.06 |
| | upper lung | 6.3 ± 9.1 | -17.7 ± 5.8 | -0.1 ± 14.1 | 0.1 |

\*p-values compare data-driven and image-based motion-corrected errors within each anatomical region.

### Influence of Respiratory Pattern and Motion Amplitude

Quantitative recovery was evaluated for the two patient-derived respiratory patterns (pattern-1 and pattern-2; **FIGURE. 1B**) and for respiratory motion amplitudes of 2 cm and 3 cm (**FIGURE. 6**). Respiratory pattern had only a minor influence on quantitative recovery. In the uncorrected datasets, SUV_mean_ errors were comparable for both respiratory patterns (pattern-1: −17.7 ± 16.3%; pattern-2: −19.2 ± 13.2%; p = 0.70). Likewise, after data-driven MoCo, residual errors remained similar for both respiratory patterns (12.6 ± 18.7% vs. 11.5 ± 8.1%; p = 0.79), and no significant respiratory-pattern dependence was observed after image-based MoCo.

**FIGURE. 6.**
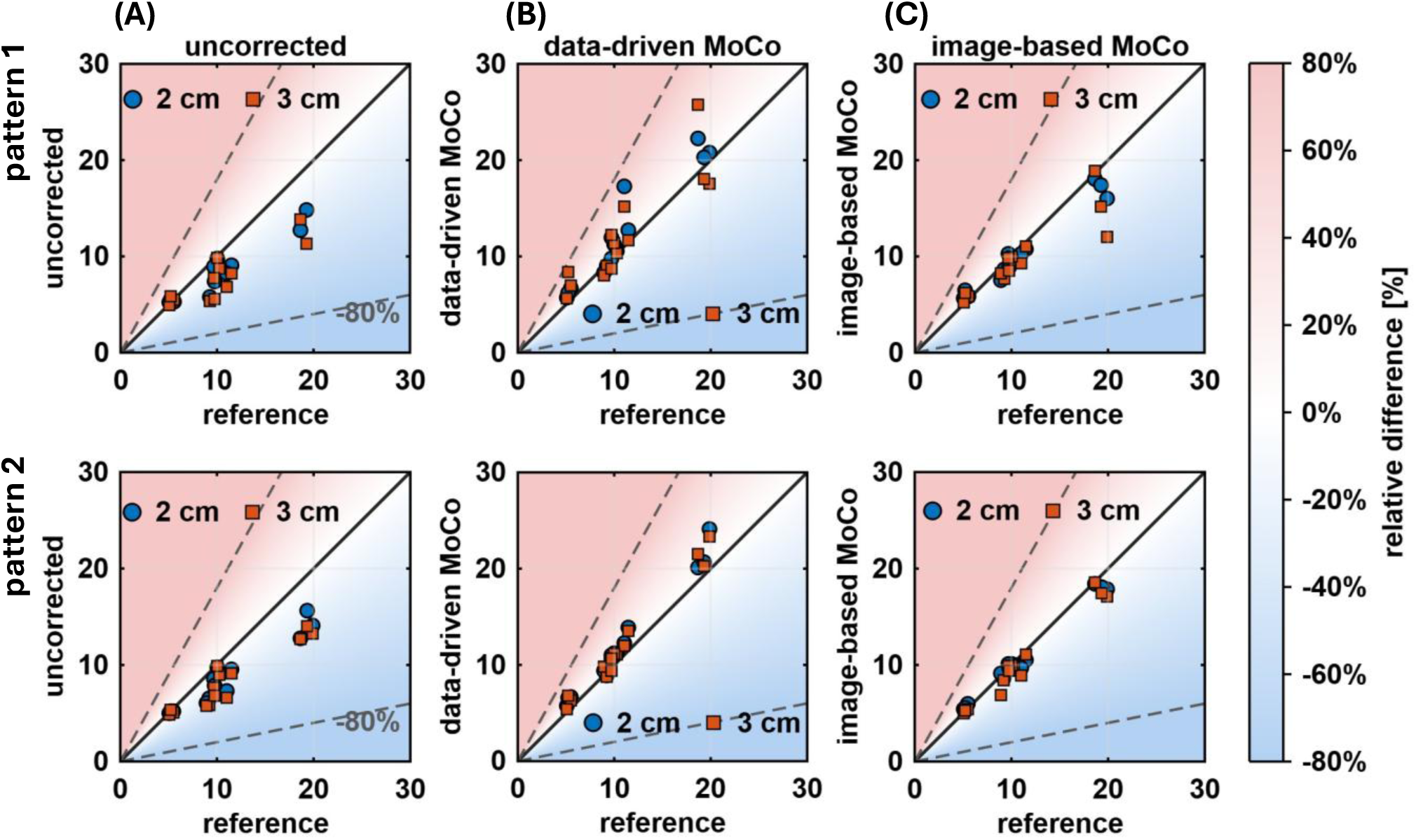
Influence of respiratory pattern and motion amplitude on SUV_mean_ recovery. Scatter plots compare lesion SUV_mean_ values of uncorrected (A), data-driven (B), and image-based MoCo (C) datasets with the corresponding motion-free reference for the two patient-derived respiratory patterns (pattern-1 and pattern-2; Figure 1B). Blue circles and orange squares indicate respiratory motion amplitudes of 2 cm and 3 cm, respectively.

In contrast, motion amplitude substantially affected quantitative recovery. Without MoCo, SUV_mean_ underestimation increased from −16.1 ± 12.8% at 2 cm to −21.0 ± 16.1% at 3 cm (p = 0.02). Data-driven MoCo remained largely independent of motion amplitude (12.6 ± 11.9% vs. 11.4 ± 16.5%; p = 0.21), whereas image- based MoCo showed increasing residual underestimation with larger motion amplitudes (−2.0 ± 8.9% vs. −6.5 ± 11.0%; p < 0.01).

## Discussion

In clinical studies, respiratory motion correction is typically evaluated by comparison with uncorrected or gated PET images because a patient-specific motion-free reference is unavailable in vivo (*19–21*). Although physical anthropomorphic motion phantoms provide valuable experimental validation (*22*,*23*), their partial-body geometries do not reproduce whole-body attenuation and scatter conditions. Digital models additionally permit efficient variation of anatomy, respiratory pattern, and motion amplitude while preserving a matched motion-free reference. By combining a validated scanner model with an anatomically realistic patient model, we established such a framework for objective assessment of quantitative accuracy. The present results demonstrate that larger correction-induced SUV recovery does not necessarily reflect greater quantitative accuracy.

Previous clinical studies consistently reported increased SUV, improved lesion conspicuity and reduced motion blurring following respiratory motion correction (*12*,*24*,*25*). Using the corresponding motion-free reference, we found that the correction strategy producing the largest apparent SUV recovery was not consistently the most accurate. Instead, quantitative performance depended on lesion location and the metric considered.

The two investigated motion correction strategies illustrate this principle. Data-driven MoCo consistently produced larger SUV increases and stronger MTV reductions in patients and simulations. However, comparison with the motion-free reference showed that this apparent advantage was confined primarily to hepatic lesions, whereas image-based MoCo remained closer to the reference for pulmonary lesions despite producing smaller correction-induced changes.

As expected, lesions adjacent to the diaphragm exhibited the largest motion-induced quantitative errors, whereas lesions farther away were considerably less affected (*1*,*2*,*12*,*26*). This regional dependence likely reflects, in part, the larger respiratory displacement near the diaphragm, consistent with the greater uncorrected error at 3- versus 2-cm motion amplitude. However, residual correction performance was not determined by amplitude alone. Data-driven MoCo was largely amplitude-independent, whereas location- and size-dependent overcorrection persisted. Additional simulations showed that overestimation after data-driven MoCo was generally greater away from the diaphragm but varied nonmonotonically with lesion position and was less pronounced for larger lesions (**Supplemental Figure 2**). Together, these observations demonstrate that correction performance is systematically influenced by lesion location, motion amplitude and size.

These findings are clinically relevant because respiratory blurring can reduce the conspicuity of small pulmonary and hepatic lesions and thereby affect lesion detection, characterization, staging, and treatment-response assessment (*4*,*19*). Ground-truth-based validation is therefore important to establish whether motion correction improves lesion depiction and quantification without introducing clinically relevant bias.

Beyond the comparison of two representative motion correction strategies, the broader contribution of this work is the presented validation framework. Although scanner-specific PET simulation frameworks have previously been developed (*27–29*), we demonstrate how they can be combined with an anatomically realistic patient model and the identical reconstruction pipeline to generate patient-like simulations incorporating patient-derived respiratory waveforms. This provides an objective framework for benchmarking respiratory motion correction algorithms under controlled yet clinically realistic conditions that cannot be achieved in patient studies alone. The same methodology can readily be transferred to other PET systems, provided similarly well-validated scanner models are available.

The proposed framework represents a controlled benchmark rather than a complete representation of clinical reality. Although multiple lesion sizes, lesion locations, respiratory amplitudes and patient-derived breathing patterns were incorporated, they cannot capture all anatomical and physiological variations encountered in clinical imaging. At the same time, the modular simulation environment readily enables extension towards different anatomies, lesion distributions, respiratory patterns and gross-body motion models, allowing increasingly realistic virtual patient populations for future algorithm development, optimization and validation.

### Conclusion

Respiratory motion correction substantially altered quantitative PET metrics clinical and simulated datasets. However, comparison with a corresponding motion-free reference demonstrated that larger correction-induced SUV recovery did not necessarily reflect greater quantitative accuracy. Data-driven motion correction most accurately recovered hepatic uptake, whereas image-based motion correction remained closer to the motion-free reference for pulmonary uptake and MTV, demonstrating that performance depends on lesion location and the metric considered.

By combining a validated scanner model with an anatomically realistic patient model, this study established a digital twin framework for objective evaluation of respiratory motion correction against a motion-free reference unavailable in clinical imaging. Because respiratory blurring may compromise small-lesion conspicuity and quantitative interpretation, such ground-truth-based validation is clinically relevant for determining whether motion correction improves lesion depiction without introducing misleading quantitative bias. As quantitative PET continues to evolve, particularly on highly sensitive LAFOV PET systems, this framework may become increasingly important for the development, optimization, and benchmarking of future PET methodologies.

## KEY POINTS

### QUESTION

Can a motion-free ground truth reveal quantitative over- and undercorrection by respiratory motion correction that cannot be identified in clinical studies?

### PERTINENT FINDINGS

In this combined clinical and simulation study of 20 patients with 135 lesions, a validated digital twin provided a corresponding motion-free reference for objective benchmarking of respiratory motion correction. It revealed region-dependent quantitative inaccuracies that were not detectable clinically, demonstrating that larger correction-induced SUV recovery did not necessarily reflect greater quantitative accuracy.

### IMPLICATIONS FOR PATIENT CARE

Ground-truth-based validation may improve quantitative PET by enabling objective optimization of respiratory motion correction for more reliable lesion depiction and quantification before clinical implementation.

## Data Availability

All data produced in the present study are available upon reasonable request to the corresponding author.

## Acknowledgements

We acknowledge Hong Phuc Vo and Christian Pommranz for their assistance during the initial setup phase of the simulation framework. We thank William Segars for insightful discussions on motion modelling in XCAT phantom and Lalith Kumar Shiyam Sundar for his support on the usage of image-based MoCo. We are also grateful to Paul Schleyer and Noah Birge for discussions on the implementation and interpretation of data-driven MoCo. We acknowledge the State of Baden-Württemberg for supporting bwHPC and the German Research Foundation for funding bwForCluster NEMO 2 (455622343).

## Supplemental Materials

**Supplemental Figure 1.**
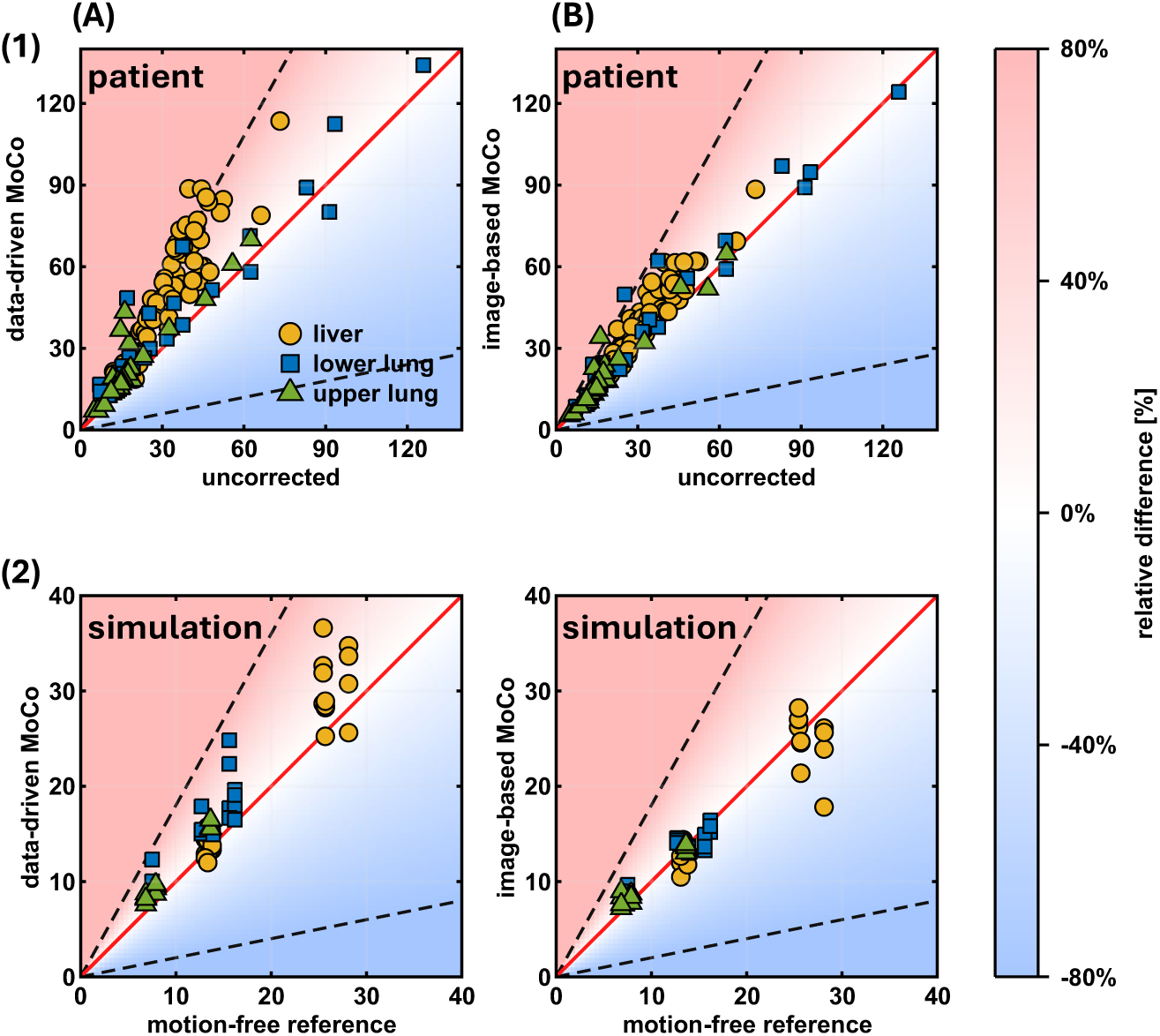
Quantitative comparison of respiratory motion correction using SUV_max_. Scatter plots compare lesion SUV_max_ values after data-driven and image-based MoCo with the corresponding uncorrected values in the clinical datasets (1) and the corresponding motion-free reference in the simulated datasets (2). Liver, lower-lung, and upper-lung lesions are indicated by circles, squares, and triangles, respectively.

**Supplemental Figure 2.**
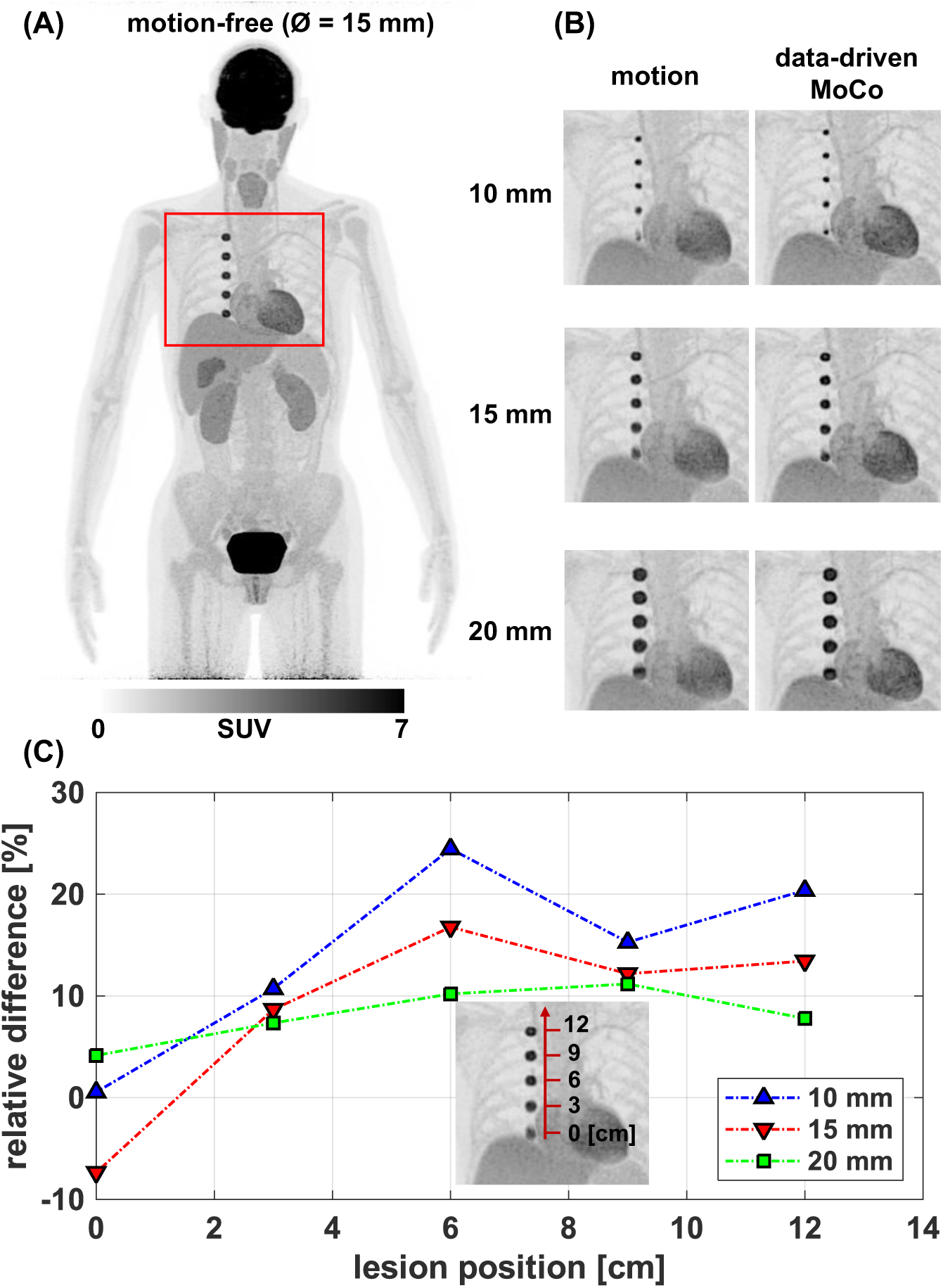
Influence of lesion location and lesion size on quantitative recovery after data- driven respiratory motion correction. (A) Maximum-intensity projection of the motion-free reference dataset containing five uniformly spaced 15-mm lung lesions. The red box indicates the enlarged region shown in (B). (B) Enlarged maximum-intensity projections of the lung lesions for diameters of 10, 15, and 20 mm under respiratory motion (2-cm amplitude) and after data-driven motion correction. (C) Relative difference in SUV_mean_ between the data-driven motion-corrected images and the corresponding motion-free reference for the five lesion positions. Results are shown separately for lesion diameters of 10, 15, and 20 mm.

**Supplemental Table 1.** Regional patient-level relative differences in SUV_mean_, SUV_max_, and MTV following data-driven and image-based respiratory motion correction. Clinical and simulated datasets are expressed relative to the corresponding uncorrected images. Values are reported as mean percentage differences (±SD).

| parameter | structure | data-driven MoCo | image-based MoCo | p-value |
| --- | --- | --- | --- | --- |
| $SUV_{mean}$<br>(patient) | liver | $48.1 \pm 18.9$ | $17.0 \pm 12.0$ | <0.01 |
| | lower lung | $32.5 \pm 21.2$ | $16.3 \pm 15.6$ | 0.06 |
| | upper lung | $28.4 \pm 32.0$ | $10.4 \pm 17.2$ | <0.01 |
| $SUV_{max}$<br>(patient) | liver | $47.0 \pm 17.9$ | $15.7 \pm 11.5$ | <0.01 |
| | lower lung | $31.8 \pm 20.4$ | $14.4 \pm 14.8$ | 0.01 |
| | upper lung | $28.9 \pm 31.8$ | $9.9 \pm 17.3$ | <0.01 |
| MTV<br>(patient) | liver | $-52.5 \pm 14.8$ | $-11.1 \pm 20.6$ | <0.01 |
| | lower lung | $-29.6 \pm 15.7$ | $-3.8 \pm 17.9$ | <0.01 |
| | upper lung | $-26.6 \pm 17.0$ | $-4.6 \pm 19.0$ | <0.01 |
| $SUV_{mean}$<br>(phantom) | liver | $55.3 \pm 16.1$ | $34.5 \pm 11.7$ | 0.01 |
| | lower lung | $45.1 \pm 29.3$ | $18.0 \pm 11.3$ | 0.06 |
| | upper lung | $17.5 \pm 6.5$ | $5.6 \pm 4.3$ | 0.06 |
| $SUV_{max}$<br>(phantom) | liver | $52.7 \pm 16.5$ | $34.8 \pm 14.5$ | 0.02 |
| | lower lung | $43.5 \pm 27.6$ | $18.0 \pm 12.8$ | 0.06 |
| | upper lung | $21.3 \pm 4.8$ | $10.6 \pm 6.4$ | 0.12 |
| MTV<br>(phantom) | liver | $-46.5 \pm 15.4$ | $-23.5 \pm 22.6$ | <0.01 |
| | lower lung | $-33.4 \pm 16.1$ | $-4.6 \pm 15.9$ | 0.02 |
| | upper lung | $-22.0 \pm 7.7$ | $-5.5 \pm 14.3$ | 0.09 |

## References

1. Liu C, Pierce LA, Alessio AM, Kinahan PE. The impact of respiratory motion on tumor quantification and delineation in static PET/CT imaging. Phys Med Biol. 2009;54:7345–7362.

2. Nehmeh SA, Erdi YE. Respiratory Motion in Positron Emission Tomography/Computed Tomography: A Review. Seminars in Nuclear Medicine. 2008;38:167–176.

3. Cherry SR, Jones T, Karp JS, Qi J, Moses WW, Badawi RD. Total-Body PET: Maximizing Sensitivity to Create New Opportunities for Clinical Research and Patient Care. J Nucl Med. 2018;59:3–12.

4. Weissensee A, Bregenzer CM, Viscione M, et al. Imaging of Small Lung Nodules on Modern SAFOV and LAFOV PET in Combination With Data-driven Motion Correction: Implications for Current Practice. Clin Nucl Med. 2026;51:300–308.

5. Artesani A, van Sluis J, van Snick JH, Providência L, Noordzij W, Tsoumpas C. Impact of patient motion on parametric PET imaging. Eur J Nucl Med Mol Imaging. January 2024.

6. Lang N, Dawood M, Büther F, Schober O, Schäfers M, Schäfers K. Organ Movement Reduction in PET/CT using Dual-Gated Listmode Acquisition. Zeitschrift für Medizinische Physik. 2006;16:93–100.

7. Ren S, Jin X, Chan C, et al. Data-driven event-by-event respiratory motion correction using TOF PET list-mode centroid of distribution. Phys Med Biol. 2017;62:4741.

8. Schleyer PJ, O’Doherty MJ, Barrington SF, Marsden PK. Retrospective data-driven respiratory gating for PET/CT. Phys Med Biol. 2009;54:1935.

9. Sundar LKS, Lassen ML, Gutschmayer S, et al. Fully Automated, Fast Motion Correction of Dynamic Whole-Body and Total-Body PET/CT Imaging Studies. Journal of Nuclear Medicine. 2023;64:1145–1153.

10. Dawood M, Buther F, Jiang X, Schafers KP. Respiratory Motion Correction in 3-D PET Data With Advanced Optical Flow Algorithms. IEEE Transactions on Medical Imaging. 2008;27:1164–1175.

11. Polycarpou I, Tsoumpas C, Marsden PK. Analysis and comparison of two methods for motion correction in PET imaging. Medical Physics. 2012;39:6474–6483.

12. Dias AH, Schleyer P, Vendelbo MH, Hjorthaug K, Gormsen LC, Munk OL. Clinical feasibility and impact of data-driven respiratory motion compensation studied in 200 whole-body 18F-FDG PET/CT scans. EJNMMI Res. 2022;12:16.

13. Nii T, Hosokawa S, Kotani T, et al. Evaluation of Data-Driven Respiration Gating in Continuous Bed Motion in Lung Lesions. Journal of Nuclear Medicine Technology. 2023;51:32–37.

14. Segars WP, Sturgeon G, Mendonca S, Grimes J, Tsui BMW. 4D XCAT phantom for multimodality imaging research. Med Phys. 2010;37:4902–4915.

15. Sarrut D, Arbor N, Baudier T, et al. The OpenGATE ecosystem for Monte Carlo simulation in medical physics. Phys Med Biol. 2022;67:184001.

16. Pommranz CM, Elmoujarkach EA, Lan W, et al. A digital twin of the Biograph Vision Quadra long axial field of view PET/CT: Monte Carlo simulation and image reconstruction framework. EJNMMI Phys. 2025;12:31.

17. Hong I, Jones J, Casey M. Ultrafast Elastic Motion Correction via Motion Deblurring. In: 2014 IEEE Nuclear Science Symposium and Medical Imaging Conference (NSS/MIC).; 2014:1–2.

18. Fedorov A, Beichel R, Kalpathy-Cramer J, et al. 3D Slicer as an image computing platform for the Quantitative Imaging Network. Magnetic Resonance Imaging. 2012;30:1323–1341.

19. Gratz M, Ruhlmann V, Umutlu L, Fenchel M, Hong I, Quick HH. Impact of respiratory motion correction on lesion visibility and quantification in thoracic PET/MR imaging. PLoS One. 2020;15:e0233209.

20. Lamare F, Bousse A, Thielemans K, et al. PET respiratory motion correction: quo vadis? Phys Med Biol. 2022;67:03TR02.

21. Meier JG, Wu CC, Cuellar SLB, et al. Evaluation of a Novel Elastic Respiratory Motion Correction Algorithm on Quantification and Image Quality in Abdominothoracic PET/CT. Journal of Nuclear Medicine. 2019;60:279–284.

22. Bolwin K, Czekalla B, Frohwein LJ, Büther F, Schäfers KP. Anthropomorphic thorax phantom for cardio- respiratory motion simulation in tomographic imaging. Phys Med Biol. 2018;63:035009.

23. Black DG, Yazdi YO, Wong J, et al. Design of an anthropomorphic PET phantom with elastic lungs and respiration modeling. Med Phys. 2021;48:4205–4217.

24. Büther F, Jones J, Seifert R, Stegger L, Schleyer P, Schäfers M. Clinical Evaluation of a Data-Driven Respiratory Gating Algorithm for Whole-Body PET with Continuous Bed Motion. J Nucl Med. 2020;61:1520–1527.

25. Overbeck N, Andersen TL, Rodell AB, et al. Device-Less Data-Driven Cardiac and Respiratory Gating Using LAFOV PET Histo Images. Diagnostics. 2024;14:2055.

26. Balfour DR, Marsden PK, Polycarpou I, Kolbitsch C, King AP. Respiratory motion correction of PET using MR-constrained PET-PET registration. BioMed Eng OnLine. 2015;14:85.

27. Merlet A, Presles B, Su K-H, et al. Validation of a discovery MI 4-ring model according to the NEMA NU 2-2018 standards: from Monte Carlo simulations to clinical-like reconstructions. EJNMMI Phys. 2024;11:13.

28. Salvadori J, Merlet A, Presles B, et al. PET digitization chain for Monte Carlo simulation in GATE. Phys Med Biol. 2024;69:165013.

29. Saaidi R, Zeghari A, El Moursli RC. Monte Carlo simulation of two Siemens Biograph PET/CT system using GATE: Image quality performance. Radiation Physics and Chemistry. 2024;218:111653.

30. Pösse S, Büther F, Mannweiler D, et al. Comparison of two elastic motion correction approaches for whole-body PET/CT: motion deblurring vs gate-to-gate motion correction. EJNMMI Phys. 2020;7:19.

31. Wang Z, Liu J, Lu D, et al. Evaluation of a motion correction algorithm in lung cancer PET/CT: Phantom validation and patient studies. Medical Physics. 2025;52:e17846.

32. Aristophanous M, Yong Y, Yap JT, et al. Evaluating FDG uptake changes between pre and post therapy respiratory gated PET scans. Radiotherapy and Oncology. 2012;102:377–382.

